# Adverse reactions to BNT162b2 mRNA COVID-19 vaccine in medical staffs with a history of allergy

**DOI:** 10.1101/2021.09.13.21263473

**Authors:** Sumito Inoue, Akira Igarashi, Keita Morikane, Osamu Hachiya, Masafumi Watanabe, Seiji Kakehata, Shinya Sato, Yoshiyuki Ueno

## Abstract

Severe acute respiratory syndrome coronavirus 2 (SARS-CoV-2, COVID-19) vaccination is progressing globally. Several adverse reactions have been reported with vaccination against COVID-19. It is unknown whether adverse reactions to COVID-19 vaccination are severe in individuals with allergies. We administered the COVID-19 vaccine to the medical staff at Yamagata University Hospital from March to August 2021. Subsequently, we conducted an online questionnaire-based survey to investigate the presence of allergy and adverse reactions after vaccination and examined the association between allergy and adverse reactions after immunization.

Responses were collected from 1586 subjects after the first vaccination and 1306 subjects after the second administration of the BNT162b2 mRNA COVID-19 vaccine. Adverse reactions included injection site pain, injection site swelling, fever, fatigue or malaise, headache, chills, nausea, muscle pain outside the injection site, and arthralgia. The frequency and severity of most adverse reactions were higher after the second vaccination compared to the first. The frequency of some adverse reactions and their severity were higher, and the duration of symptoms was longer in subjects with allergies than in subjects without allergies. Although several participants visited the emergency room for treatment after the first and second vaccinations, nobody was diagnosed with anaphylaxis.

Given the serious consequence of COVID-19 and the reported high efficacy of this vaccine against this disease, we conclude that vaccination of allergic individuals is generally recommended.

## Introduction

As the severe acute respiratory syndrome coronavirus 2 (SARS-CoV-2, COVID-19) pandemic continues to spread worldwide, COVID-19 vaccination is one of the solutions to control it. BNT162b2 mRNA COVID-19 vaccine (Pfizer-BioNTech) is the first available mRNA vaccine in Japan in February 2021, which was initially administered to healthcare workers (1). Currently, several types of COVID-19 vaccines are available, and many people are being vaccinated (1, 2, 3). Although the efficacy of COVID-19 vaccination varies depending on the type of vaccine, the BNT162b2 has been reported to be 95% effective in preventing symptomatic COVID-19 infection in vaccinated individuals, indicating its high efficacy (1). Effective medications against COVID-19 include dexamethasone (4, 5), an antiviral agent remdesivir (6), and Janus kinase (JAK) inhibitor baricitinib (7), all of which are currently available in Japan for COVID-19 treatment. Although various other drugs are candidates for COVID-19 treatment, negative data on their efficacy, including those currently available, have been reported; hence, no reliable treatment is available (8).

Therefore, suppression of the COVID-19 pandemic by vaccination is considered important. However, there is a risk for adverse reactions to vaccination, and various adverse reactions have been reported with the COVID-19 vaccine (9). Adverse reactions such as pain at the vaccination site or fever are the main adverse reactions that have been reported. A particular problem with adverse reactions is allergic symptoms, and if anaphylaxis develops, there is a risk of life-threatening consequences (1-3). Since most people are receiving the COVID-19 vaccine for the first time, it is largely unknown whether the vaccination will cause adverse reactions, such as allergy or severe anaphylaxis. Particularly, it is difficult to determine whether individuals with allergies can receive the COVID-19 vaccine. However, to verify the safety of COVID-19 vaccines, individuals with a history of allergy to vaccine components were excluded from clinical trials. As a result, very few studies have examined the safety of COVID-19 vaccination in people with allergies.

At Yamagata University Hospital, Japan, BNT162b2 was administered to hospital staff and students. After vaccination, we conducted a questionnaire survey to find out their allergy history and adverse reactions after immunization. By analyzing these data, we aimed to verify the safety of the COVID-19 vaccine in people with a history of allergy.

## Materials and Methods

### Study Design

The questionnaire survey was administered to the medical staff of Yamagata University Hospital and staff and medical students of Yamagata University Faculty of Medicine who received the BNT162b2 mRNA COVID-19 vaccine (Pfizer-BioNTech) from March 3, 2021 to August 27, 2021. After the first and second vaccination, a paper containing an internet link to the questionnaire was distributed. The questionnaire was designed using the free web-based Google Forms software. We asked the subjects their gender, age, occupation, history of allergy to food and/or medicine, history of allergic diseases, history of anaphylaxis, and history of adverse reactions to vaccination. As for adverse reactions after the immunization, the subjects were asked about injection site pain, injection site swelling, fever, fatigue or malaise, headache, chills, nausea, muscle pain outside the injection site, and arthralgia, as well as the timing of adverse reactions appearance and duration and degree of symptoms. The Institutional Ethics Committee of the Yamagata University Faculty of Medicine approved this study (approval number; 2021-130, approval date: June 29, 2021). The opt-out method was used to obtain informed consent, which is available on our website. Patients who refused to participate were excluded from the study. The individuals participated anonymously.

### Statistical Analysis

All classified variables are presented as numbers and percentages. The differences between groups were evaluated using the chi-squared test. Significance was inferred for p values of <0.05. Statistical analyses were performed using JMP version 11.0 software (SAS Institute, Cary, NC, USA).

## Results

There were 1586 questionnaires returned from individuals who had received the first vaccination of BNT162b2, and 1306 questionnaires were returned from individuals who had received the second vaccination after approximately 3 weeks. Table 1 shows the profiles and allergy histories of individuals. Those who received the first vaccination were 522 (32.9%) male and 1064 (67.1%) female. By age group, 546 participants (34.4%) were in their 20s, 402 (25.3%) were in their 30s, 336 (21.2%) were in their 40s, 220 (13.9%) were in their 50s, and 82 (5.2%) were in their 60s or older. Of the participants, 193 (12.2%) had a history of allergy to food and/or medicine. There were 698 (44.0%) participants with allergic diseases, such as rhinitis and bronchial asthma, 27 (1.7%) had a history of anaphylaxis, and 90 (5.7%) had a history of adverse reactions after vaccination. The profiles were similar to those after the second vaccination.

**Table 1.** Profile of subjects vaccinated with BNT162b2 mRNA COVID-19 vaccine

|  | First vaccination (1586) | Second vaccination (1306) |
| --- | --- | --- |
| Male / Female | 522 / 1064 | 388 / 918 |
| Type of Job |  |  |
| Doctor | 320 (20.3%) | 216 (16.6%) |
| Nurse | 558 (35.3%) | 509 (39.0%) |
| Technician | 152 (9.6%) | 123 (9.4%) |
| Clerk | 371 (23.4%) | 299 (22.9%) |
| Student | 179 (11.3%) | 157 (12.0%) |
| Unknown (no answer) | 6 (3.8%) | 2 (0.2%) |
| Age |  |  |
| 20-29 | 546 (34.4%) | 427 (32.7%) |
| 30-39 | 402 (25.3%) | 321 (24.6%) |
| 40-49 | 336 (21.2%) | 288 (22.1%) |
| 50-59 | 220 (13.9%) | 197 (15.1%) |
| 60- | 82 (5.2%) | 73 (5.6%) |
| History of allergy (food and/or medicine) | 193 (12.2%) | 160 (12.3%) |
| History of allergic diseases | 698 (44.0%) | 564 (43.2%) |
| History of anaphylaxis | 27 (1.7%) | 13 (1.0%) |
| History of adverse response to vaccination | 90 (5.7%) | 91 (7.0%) |

The frequency of adverse reactions after first and second vaccinations is shown in Figure 1. Adverse reactions other than pain at the injection site occurred more frequently after the second vaccination. Table 2 summarizes the onset of adverse reactions, duration of symptoms, and severity of symptoms after the first and second vaccinations. In this study, adverse reactions after vaccination other than fever were defined as follows: moderate adverse reactions were those that interfered with daily life, and severe adverse reactions were those that required medical treatment. Moderate fever and severe fever were defined as a body temperature of 38°C or higher, and body temperature of 39°C or higher, respectively. For all adverse reactions, the frequency of symptoms lasting more than 2 days was high. The frequency of moderate or severe symptoms was also high.

**Figure 1.**
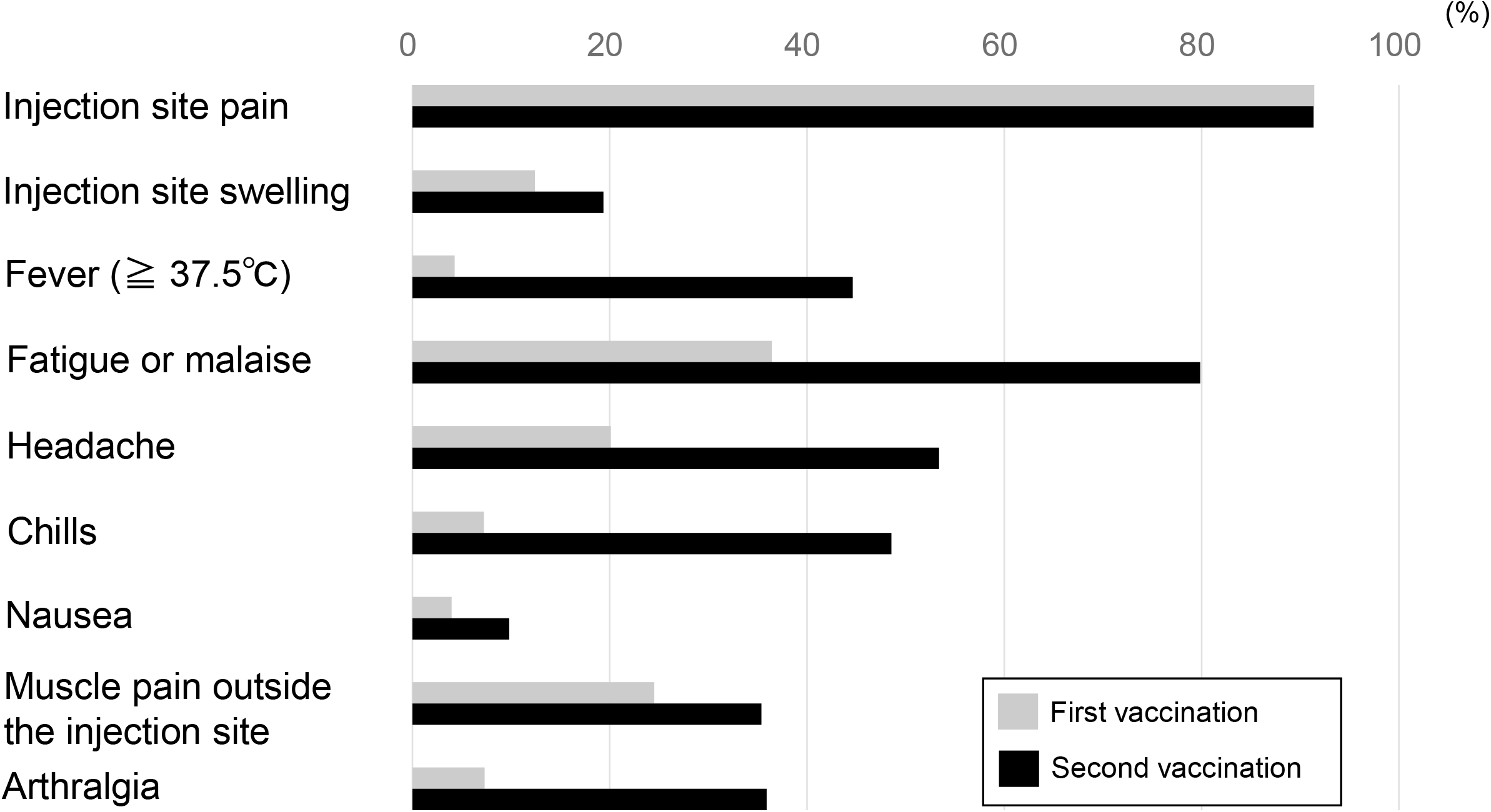
Frequency of adverse reactions after the first and second vaccinations. Adverse reactions other than pain at the injection site occurred more frequently after the second vaccination than after the first.

**Table 2.** The onset of adverse reactions, duration of symptoms, and severity of symptoms after the first and second vaccination

|  |  | Number of cases (%) | Onset on the day of vaccination (%) | Continued for more than 2 days (%) | Moderate or severe symptoms (%) |
| --- | --- | --- | --- | --- | --- |
| Injection site pain | After first vaccination | 1450 (91.4) | 957 (60.3) | 981 (61.9) | 164 (10.3) |
|  | After second vaccination | 1193 (91.3) | 807 (61.8) | 896 (68.6) | 437 (33.5) |
| Injection site swelling | After first vaccination | 197 (12.4) | 100 (6.3) | 106 (6.7) | 8 (0.5) |
|  | After second vaccination | 253 (19.4) | 104 (8.0) | 176 (13.5) | 42 (3.2) |
| Fever ($\geq 37.5$ °C) | After first vaccination | 68 (4.3) | 18 (1.1) | 16 (1.0) | 23 (1.5) |
|  | After second vaccination | 583 (44.6) | 134 (10.3) | 185 (14.2) | 322 (24.7) |
| Fatigue or malaise | After first vaccination | 578 (36.4) | 272 (17.2) | 246 (15.5) | 68 (4.3) |
|  | After second vaccination | 1043 (79.9) | 315 (24.1) | 573 (43.9) | 561 (43.0) |
| Headache | After first vaccination | 319 (20.1) | 151 (9.5) | 107 (6.7) | 123 (7.8) |
|  | After second vaccination | 697 (53.4) | 200 (15.3) | 364 (27.9) | 493 (37.7) |
| Chills | After first vaccination | 115 (7.3) | 52 (3.3) | 30 (1.9) | 28 (1.8) |
|  | After second vaccination | 634 (48.5) | 224 (17.2) | 219 (16.8) | 375 (28.7) |
| Nausea | After first vaccination | 63 (4.0) | 42 (2.6) | 14 (0.9) | 10 (0.6) |
|  | After second vaccination | 128 (9.8) | 41 (3.1) | 43 (3.3) | 35 (2.7) |
| Muscle pain outside the injection site | After first vaccination | 389 (24.5) | 210 (13.2) | 262 (16.5) | 56 (3.5) |
|  | After second vaccination | 462 (35.4) | 179 (13.7) | 278 (21.3) | 198 (15.2) |
| Arthralgia | After first vaccination | 116 (7.3) | 44 (2.8) | 50 (3.2) | 16 (1.0) |
|  | After second vaccination | 469 (35.9) | 107 (8.2) | 221 (16.9) | 264 (20.2) |

Table 3 summarizes the frequency of adverse reactions by gender and age. The frequency of fatigue or malaise, headache, chills, nausea, and muscle pain outside the injection site after the first vaccination was significantly higher in females than in males. After the second vaccination, the frequency of fever, fatigue or malaise, headache, chills, nausea, muscle pain outside the injection site, and arthralgia was significantly higher in females than that in males. No adverse reactions occurred with a significantly higher incidence in males. In the analysis by age, the frequency of injection site pain, injection site swelling, fever, and nausea after the first vaccination was significantly higher in younger people. The frequency of injection site pain, fever, fatigue or malaise, headache, chills, nausea, and muscle pain outside the injection site after the second vaccination was significantly higher in the younger age group.

**Table 3.**
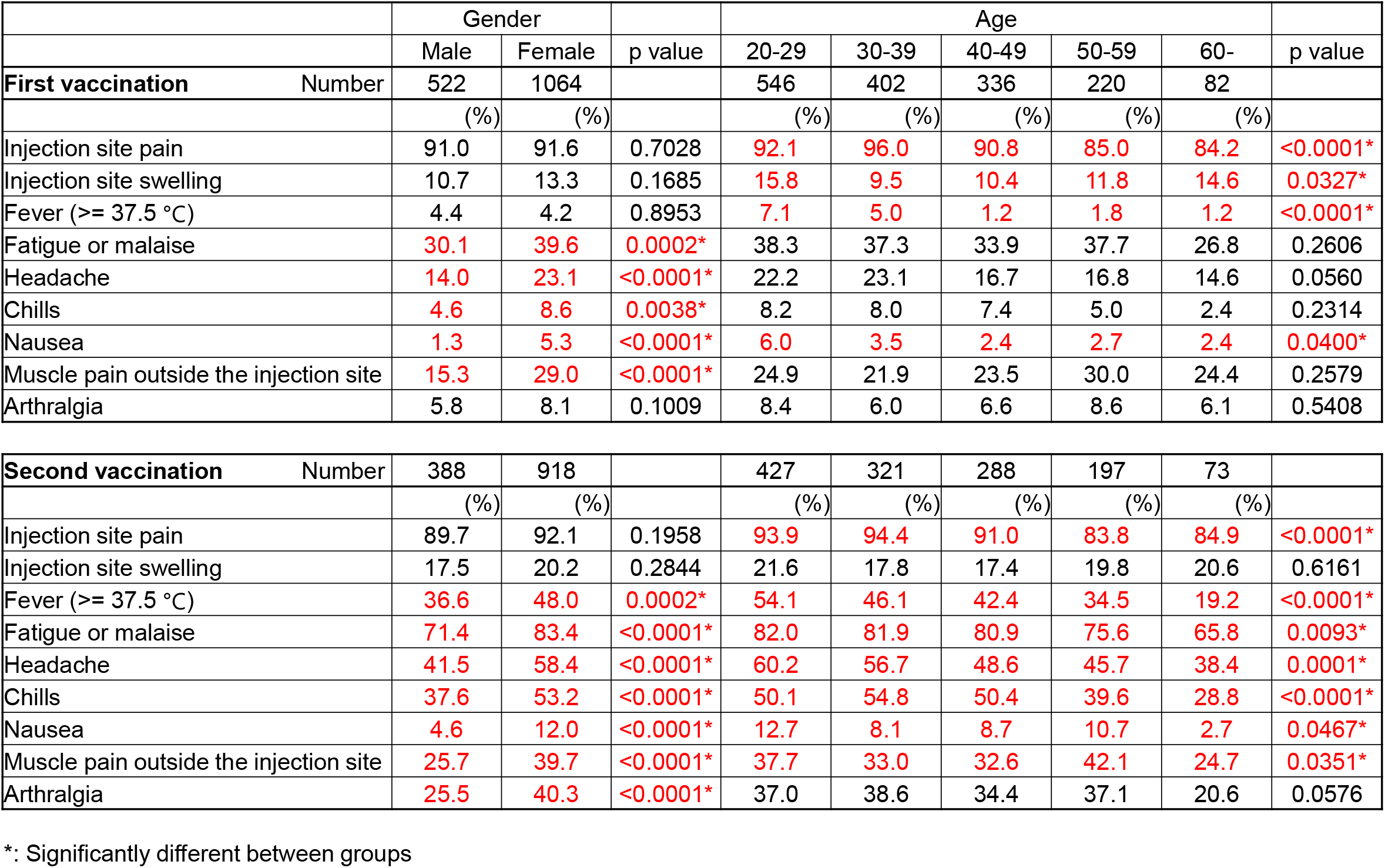
The frequency of adverse reactions by gender and age.

Table 4 summarizes the frequency of adverse reactions by allergic status. Those who have a history of allergy to food and/or medicine had a significantly higher incidence of fatigue or malaise, headache, chills, nausea, and arthralgia after the first vaccination and a higher incidence of headache, nausea, muscle pain outside the injection site, and arthralgia after the second vaccination than those without a history of allergy to food and/or medicine. Those who have a history of allergic diseases, such as bronchial asthma and allergic rhinitis, had a significantly higher incidence of injection site pain, injection site swelling, fever, fatigue or malaise, headache, nausea, and arthralgia after the first vaccination, and a higher incidence of injection site swelling and nausea after the second vaccination than those without a history of allergic diseases. Those who have a history of anaphylaxis had a significantly higher incidence of fatigue or malaise after the first vaccination and a lower incidence of headache after the second vaccination than those without a history of anaphylaxis. Those who have a history of adverse reactions after vaccination had a significantly higher incidence of injection site swelling, fever, fatigue or malaise, headache, and chills after the first vaccination and a higher incidence of injection site pain and injection site swelling, nausea, and arthralgia after the second vaccination than those without a history of adverse reactions after vaccination.

**Table 4.**
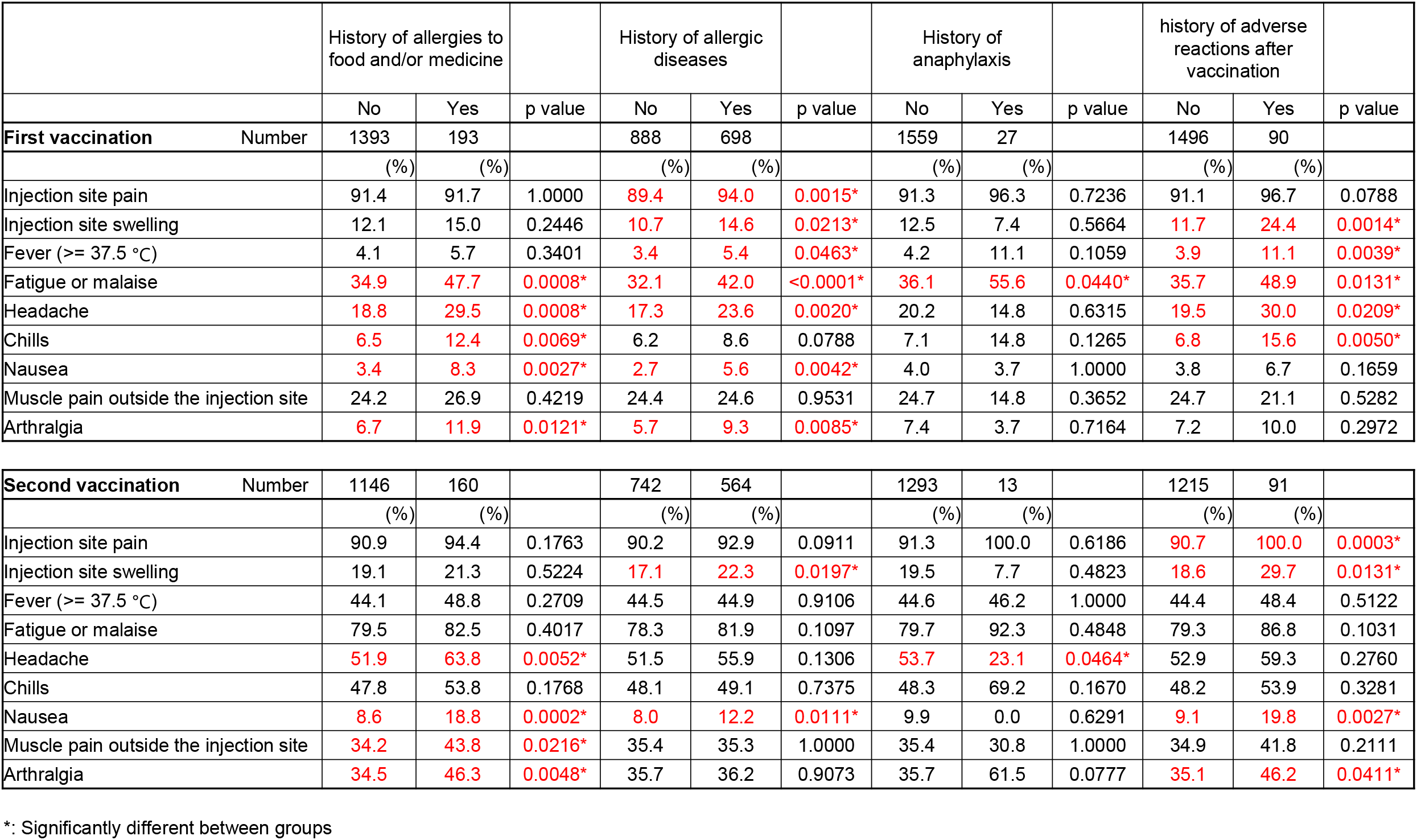
The frequency of adverse reactions by allergic status.

Table 5 shows whether individuals with allergies continued to have adverse reactions after vaccination for a long period of time. The frequency of fatigue or malaise after the first and second vaccinations and headache and chills after the second vaccination, lasting more than two days, was significantly higher among those with a history of allergy to food and/or medicine than those without. The frequency of fever after the first vaccination and injection site pain after the second vaccination, lasting more than two days, was significantly higher among those with a history of allergy than those without. The frequency of adverse reactions lasting more than 2 days after vaccination did not differ with the presence or absence of a history of anaphylaxis. The frequency of fatigue or malaise and headache after the second vaccination, lasting more than two days, was significantly higher among those with a history of adverse reactions after vaccination than those without.

**Table 5.**
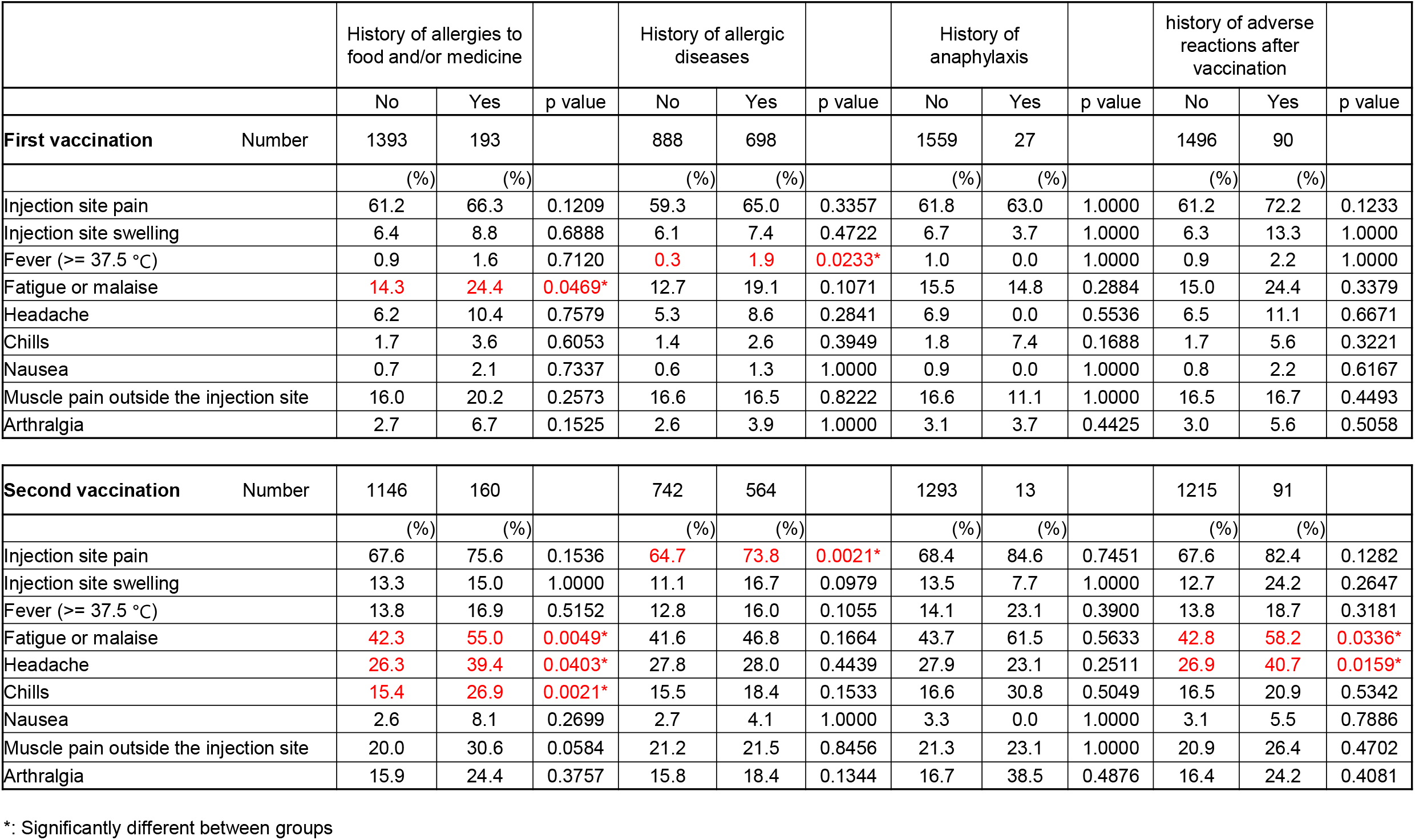
The frequency of adverse reactions continued more than 2 days

Table 6 demonstrates whether the frequency of moderate to severe adverse reactions after vaccination differs, according to the presence or absence of allergy. Individuals with a history of allergy to food and/or medicine had significantly a higher incidence of moderate to severe arthralgia after the first vaccination and muscle pain outside the injection site after second vaccination than those without. The frequency of moderate to severe adverse reactions after vaccination did not differ with the presence or absence of a history of allergic diseases or anaphylaxis. Individuals with a history of adverse reactions after vaccination had a significantly higher incidence of moderate to severe fatigue or malaise after the first vaccination and injection site pain and chills after the second vaccination than those without. After the first and second vaccinations, a total of four subjects (one male and three females in their 20s to 40s) visited the emergency room for treatment, but none were diagnosed with anaphylaxis. One of them had a food allergy and allergic rhinitis while the other three had no allergy. No other life-threatening adverse reactions or deaths were reported.

**Table 6.**
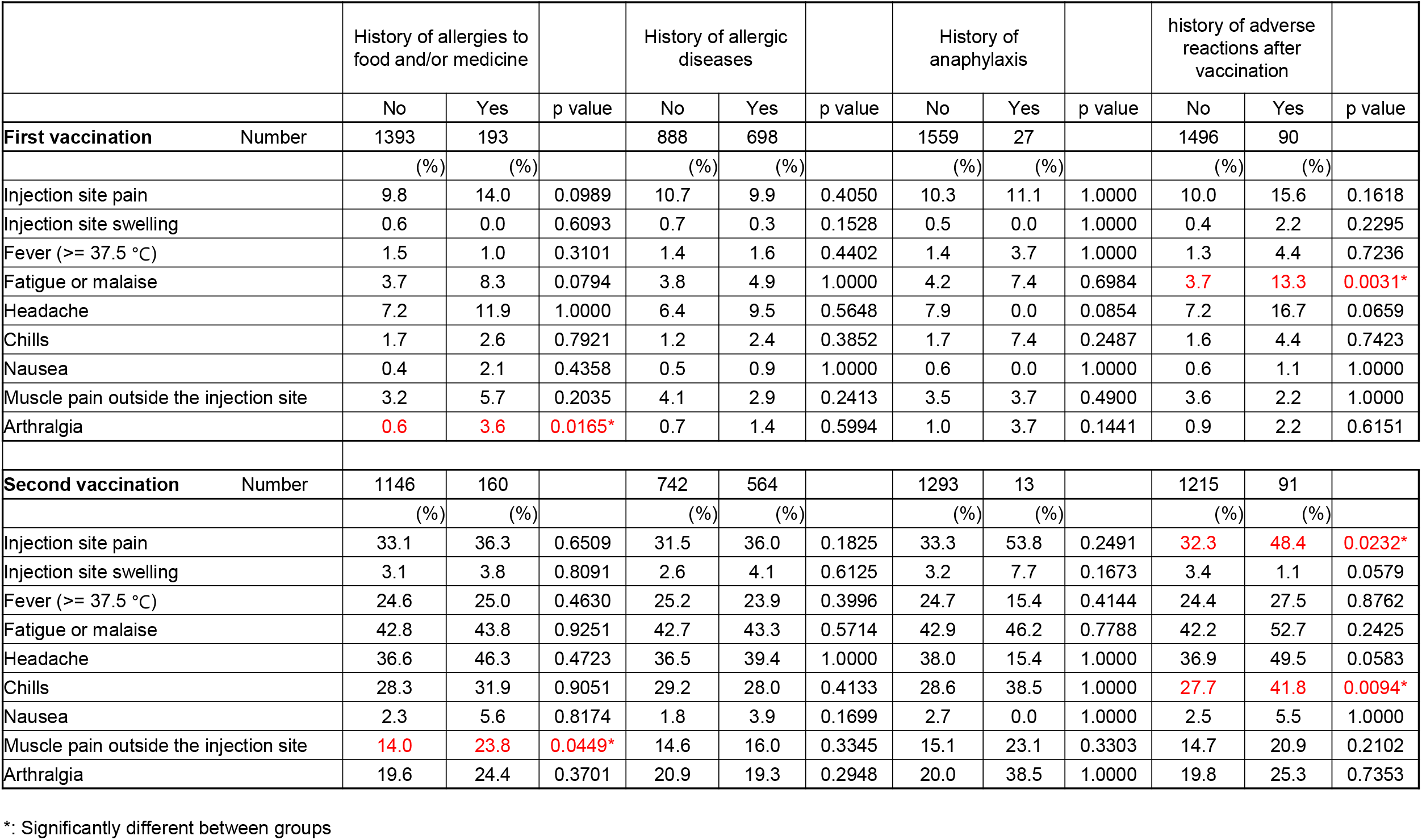
The frequency of moderate to severe reactions (for fever, body temperature of 38°C or higher, and for other adverse reactions, those that interfere with daily life or require medical treatment)

## Discussion

We administered the BNT162b2 to medical staff and investigated post-vaccination adverse reactions. The results showed that the frequency of adverse reactions after the COVID-19 vaccination was higher, the duration of symptoms was longer, and the symptoms were more severe after the second vaccination than after the first. Females and younger individuals experienced adverse reactions at a higher rate than males and the elderly, respectively. These results were consistent with previous reports (10). In our present analysis, we examined adverse reactions after COVID-19 vaccination in participants with allergies. Those who have a history of allergy to food and/or medicine, allergic diseases such as bronchial asthma or allergic rhinitis, a history of anaphylaxis or adverse reactions after vaccination had a significantly higher frequency of some adverse reactions compared to those without. Some adverse reactions showed a higher frequency of longer symptom duration after vaccination and a higher frequency of moderate or severe adverse reactions, but the differences were not outstandingly large. Additionally, no serious life-threatening allergic events were experienced after the BNT162b2 vaccination at our hospital.

In a previous report, the COVID-19 vaccination of hospital staff was performed, and the frequency of adverse reactions was examined according to the presence or absence of allergy (11). They found that subjects with a history of allergy had a significantly higher incidence of adverse reactions after vaccination than those without. They concluded that the BNT162b2 is less well tolerated in allergic individuals than in non-allergic participants, but the adverse reactions that occurred were mild and did not interfere with the successful completion of vaccination. Our results and conclusions are comparable, but our study is valuable since we not only investigated the presence or absence of allergy but also a detailed history of anaphylaxis and previous histories of adverse reactions to vaccination. Furthermore, our data are from the Japanese population. Therefore, we believe that our findings will be useful information for Japanese people who have a history of allergy to food and/or medicine, history of allergic diseases, history of anaphylaxis, and history of adverse reactions to vaccination.

There are several limitations to our study. The first limitation is that the survey was an internet-based questionnaire, and symptoms and allergy history were self-reported. Particularly, it is unclear whether the diagnosis of allergic diseases is correct. Secondly, not many subjects had a history of anaphylaxis or adverse reactions to vaccinations; thus, the statistical reliability of the study may not be high. Thirdly, it is possible that some of the staff did not receive the vaccine because of a history of allergy. Therefore, it is possible that some people with a history of allergies may not have participated in this study. It is expected that large-scale and detailed data on adverse reactions to COVID-19 vaccination will be accumulated in the future, and the advantages and disadvantages of vaccination, especially for people with a history of allergy, will be verified.

In conclusion, although the tolerance of BNT162b2 was worse in individuals with allergies than those without, no serious adverse reactions such as anaphylaxis or death were observed. For those who are hesitant about COVID-19 vaccination because of allergy, data from a large adverse reaction study can be expected to be very helpful. We hope that the results of this study will be used to successfully complete the vaccination.

## Supporting information

ICMJE DISCLOSURE FORM

ICMJE DISCLOSURE FORM

ICMJE DISCLOSURE FORM

ICMJE DISCLOSURE FORM

ICMJE DISCLOSURE FORM

ICMJE DISCLOSURE FORM

ICMJE DISCLOSURE FORM

ICMJE DISCLOSURE FORM

## Data Availability

All data will not be made public in principle because some parts contain personal information of the subjects.
For ethical reasons, data that is not suitable for public disclosure may be made available only upon request.

## Acknowledgment

The authors would like to thank all the health care professionals involved in COVID-19 treatment at Yamagata University Hospital. The authors would like to thank Enago (www.enago.jp) for the English language review.

